# Hantavirus Disease in Uruguay: Trends and Mortality Before and During the COVID-19 Pandemic

**DOI:** 10.64898/2026.06.10.26355375

**Authors:** Zelika Criscuolo, Blanco Mila, Federico Ferrara, Karen Ciaccio, Leandro Gómez Carassale, María González Reyes, Bruno Machado Rivero, Federico Sosa Dias, Jorge Facal Castro

## Abstract

**Introduction:** Hantavirus disease is an emerging and potentially severe zoonosis of global distribution. In Uruguay, it is transmitted by rodents inhabiting peridomestic, suburban, and rural areas. Global incidence is estimated at 150,000 to 200,000 cases per year, with up to 300 annual cases in the Americas. Since 1997, Uruguay’s Ministry of Public Health (MPH) has monitored Hantavirus cardiopulmonary syndrome (HCPS), the most common clinical presentation in the region. By 2019, a total of 271 cases had been identified in the country, with an estimated mortality rate of nearly 50%.

**Objectives:** To describe the clinical, epidemiological, and occupational characteristics of patients with Hantavirus disease in Uruguay during the pre-pandemic (2018-2019) and pandemic (2020-2021) periods.

**Methods:** A descriptive, cross-sectional, observational study was conducted, including all serologically confirmed cases of Hantavirus infection reported to the MPH between 2018 and 2021. Clinical and demographic data were extracted from the mandatory reporting form for zoonotic diseases. Incidence and case fatality rates were calculated, and factors associated with fatal outcomes were analyzed.

**Results:** A total of 58 confirmed cases were identified between 2018 and 2021. Most patients were male (62%), with a mean age of 36.5 years (SD 16). A decline in incidence was observed during 2020-2021, with no significant change in case fatality. Direct rodent exposure was the most frequently associated risk factor. Montevideo and Canelones were the most affected departments. Renal and pulmonary involvement were significantly associated with mortality.

**Conclusion:** Hantavirus remains a relevant public health concern in Uruguay. Although a decrease in incidence was observed during the COVID-19 pandemic years, case fatality rates remained high. The findings underscore the need for sustained surveillance and early recognition, particularly in urbanizing regions.

## Introduction

Hantavirus disease is a potentially severe zoonosis caused by viruses of the genus *Hantavirus* (family *Bunyaviridae*), with at least 28 species known to infect humans [1]. Global distribution follows that of rodent reservoirs, with an estimated annual incidence of 150,000–200,000 cases worldwide and up to 300 cases in the Americas, where at least 13 countries have endemic areas [2].

In Uruguay, the Ministry of Public Health (MPH) initiated national surveillance for hantavirus cardiopulmonary syndrome (HCPS) in 1997. HCPS, the only clinical form reported in the country, is transmitted to humans mainly by inhalation of aerosols from excreta of chronically infected rodents, particularly the field mouse (*Akodon spp*.) and the long-tailed mouse (*Oligoryzomys spp*.), found in rural and peri-urban environments [3,6].

Environmental and occupational conditions influencing hantavirus transmission have been described in neighboring Argentina, where factors such as rodent habitat proximity and human activities in rural areas are associated with increased risk [4]. The disease has three clinical phases—prodromal, cardiopulmonary, and convalescent—with a reported mortality of 40–50% [1,3].

No specific antiviral therapy or vaccine approved by the U.S. Food and Drug Administration is available; management remains supportive [5]. Recent national reports highlight the need for updated epidemiological data, particularly in the context of the COVID-19 pandemic [6]. This study aims to describe clinical, epidemiological, and occupational features of hantavirus cases in Uruguay, comparing the pre-pandemic period with the first two years of the pandemic.

## Objectives

To describe the clinical and epidemiological characteristics of patients with serologically confirmed hantavirus disease reported to the Ministry of Public Health (MPH) of Uruguay during the pre-pandemic period (2018–2019) and the first two years of the SARS-CoV-2 pandemic (2020–2021). To estimate the incidence and case fatality rate of the disease, and to describe associations between demographic, clinical, and laboratory variables and case fatality.

## Methods

A descriptive, cross-sectional observational study was conducted at the Academic Medical Unit “1” of Hospital Maciel, Montevideo, Uruguay, in collaboration with the Department of Epidemiology of the Ministry of Public Health (MPH). All variables contained in the “Zoonotic Disease Notification Form” for patients reported and serologically confirmed with hantavirus infection were requested from the Department of Epidemiological Surveillance of the MPH, using the “Epidemiological Information Request Form” available on the MPH website. Data were received in an anonymized Excel file, in compliance with the principles of ethics in clinical research and personal data protection legislation.

All cases reported to the MPH with serologically confirmed hantavirus infection between 2018 and 2021 in Uruguay were included.

The variables analyzed were **demographic and occupational:** age, sex, cleaning of sheds or storage facilities, contact with rodents, visits to wild areas, risk exposure, place of residence, and department. Except for the last two, which were qualitative nominal variables, all others were dichotomous. **Clinical and laboratory variables:** fever, jaundice, pneumonia or pneumonitis, digestive symptoms, hemorrhagic syndrome, headache, myalgia and/or arthralgia, abnormal chest radiography, leukocytosis, thrombocytopenia, and renal failure. **Outcome variables:** hospitalization and death.

Normality of data was assessed. Quantitative variables were presented as mean and standard deviation (SD), and categorical variables as absolute frequency and relative frequency (percentage). Analyses were performed using available data. For quantitative variables with missing values, mean imputation was applied; categorical variables with missing entries were excluded from percentage calculations.

Incidence was calculated as the total number of new hantavirus cases per year divided by the population of Uruguay, multiplied by 1,000,000 inhabitants. Following the “Updated Report: Hantavirus Situation in Uruguay, January 2019,” the population was divided into two regions—south and north of the Río Negro—and incidence was calculated for each region. Population data for Uruguay and each department were obtained from the 2011 National Census.

Case fatality rate was calculated as the number of hantavirus-related deaths per year divided by the number of confirmed hantavirus cases in that year, multiplied by 100. Associations between variables and case fatality were evaluated using the Chi-square or Fisher’s exact test for categorical variables, and Student’s t-test or Mann–Whitney U test for continuous variables, as appropriate. The odds ratio (OR) of various clinical variables for death was estimated using a univariate logistic regression model, with 95% confidence intervals (95% CI).

The pre-pandemic period was defined as January 1, 2018, to March 13, 2020, when the first confirmed SARS-CoV-2 case was reported in Uruguay.

Statistical analysis was performed using IBM SPSS software. All study data were handled with strict confidentiality. The protocol was reviewed and approved by the Ethics Committee of Hospital Maciel.

## Results

### General characteristics

Between 2018 and 2021, 58 hantavirus cases were confirmed in Uruguay. Most patients were male (62.1%), with a mean age of 36.5 years. Over half of the cases (55.2%) occurred in individuals aged 15-39 years, followed by 40-60 years (29.3%). The vast majority of cases were reported in departments south of the Río Negro (96.6%), with Montevideo accounting for 32.8% of cases, followed by Canelones (22.4%) and Colonia (12.1%) (Figure 1). Regarding place of residence, 36.2% of patients lived in urban areas, 27.6% in suburban areas, and 19% in rural areas.

**Figure 1.**
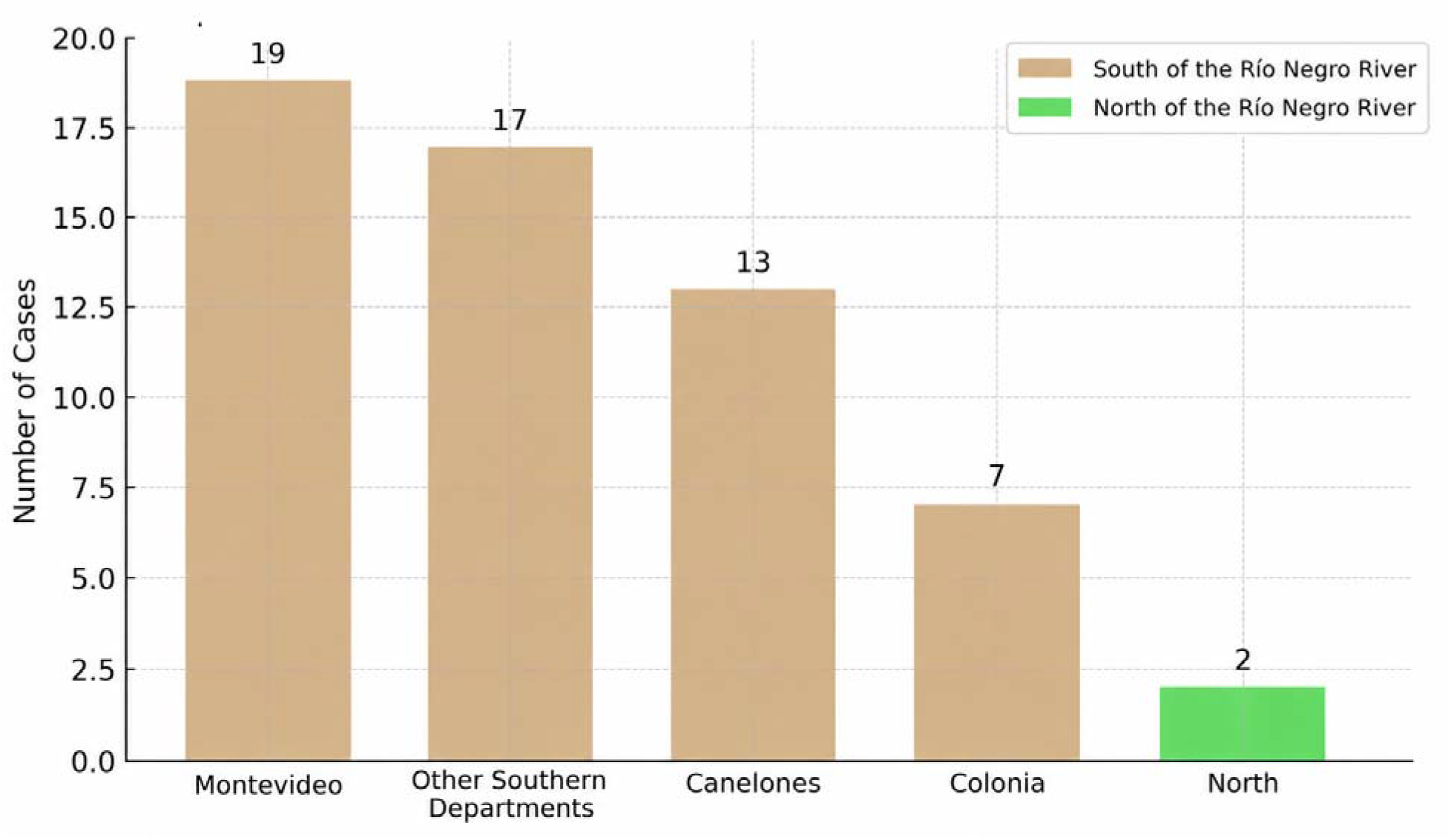
Distribution of Hantavirus cases by Uruguay department, 2018-2021.

### Incidence and case fatality rates

The annual incidence per 1,000,000 inhabitants was 6.7 in 2018, 5.2 in 2019, 3.4 in 2020, and 2.4 in 2021 (Figure 2). The case fatality rate was 27.3% in 2018, 17.6% in 2019, 27.3% in 2020, and 37.5% in 2021. When comparing pre-pandemic and pandemic periods, incidence rates were 13.69 and 3.95 per 1,000,000 inhabitants, respectively, with case fatality rates of 26.7% and 23.1%.

**Figure 2.**
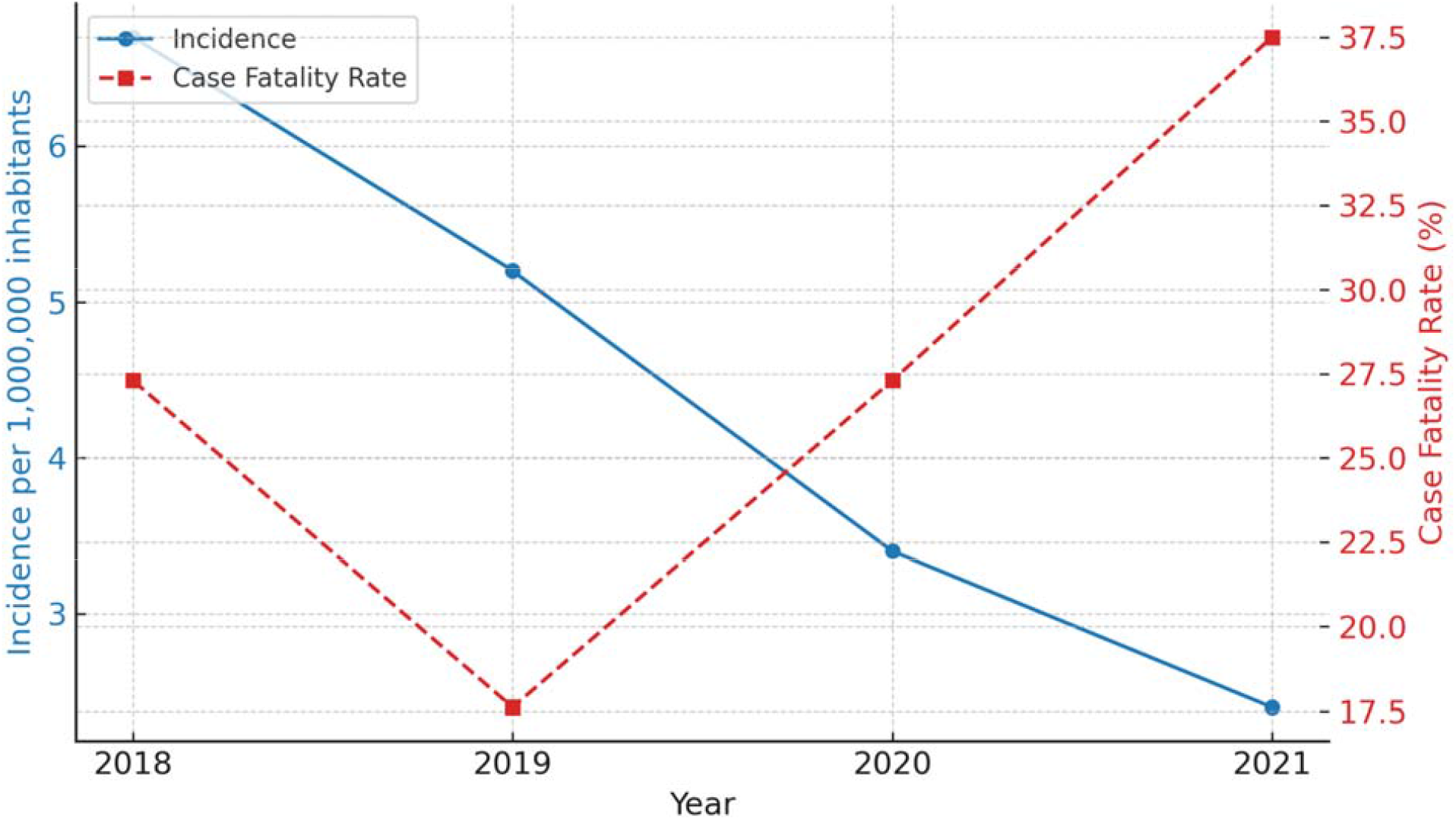
Trends in Hantavirus incidence and case fatality rate in Uruguay (2018-2021).

In the south of the Río Negro, incidence was 7.3 per 1,000,000 inhabitants in 2018, 5.9 in 2019, 4.0 in 2020, and 2.93 in 2021, while in the north, it was 1.8 in 2018 and 2019, and 0 in 2020 and 2021. The hospitalization rate was 77.6% (n=45; 95% CI 66.9-88.3), with a case fatality rate among hospitalized patients of 31.1%.

### Risk factors and clinical features

The most frequently reported risk factor was contact with rodents (39.7%), followed by cleaning of sheds (31%) and visiting wild areas (20.7%) (figure 3). Among the 58 cases, 87.9% (n=51) presented with fever, 72.4% (n=42) with myalgia and/or arthralgia, and 62.1% (n=34) with headache. Laboratory findings showed thrombocytopenia and renal involvement in up to 50% of cases, and hepatic involvement in 38% (Figure 4 and 5).

**Figure 3.**
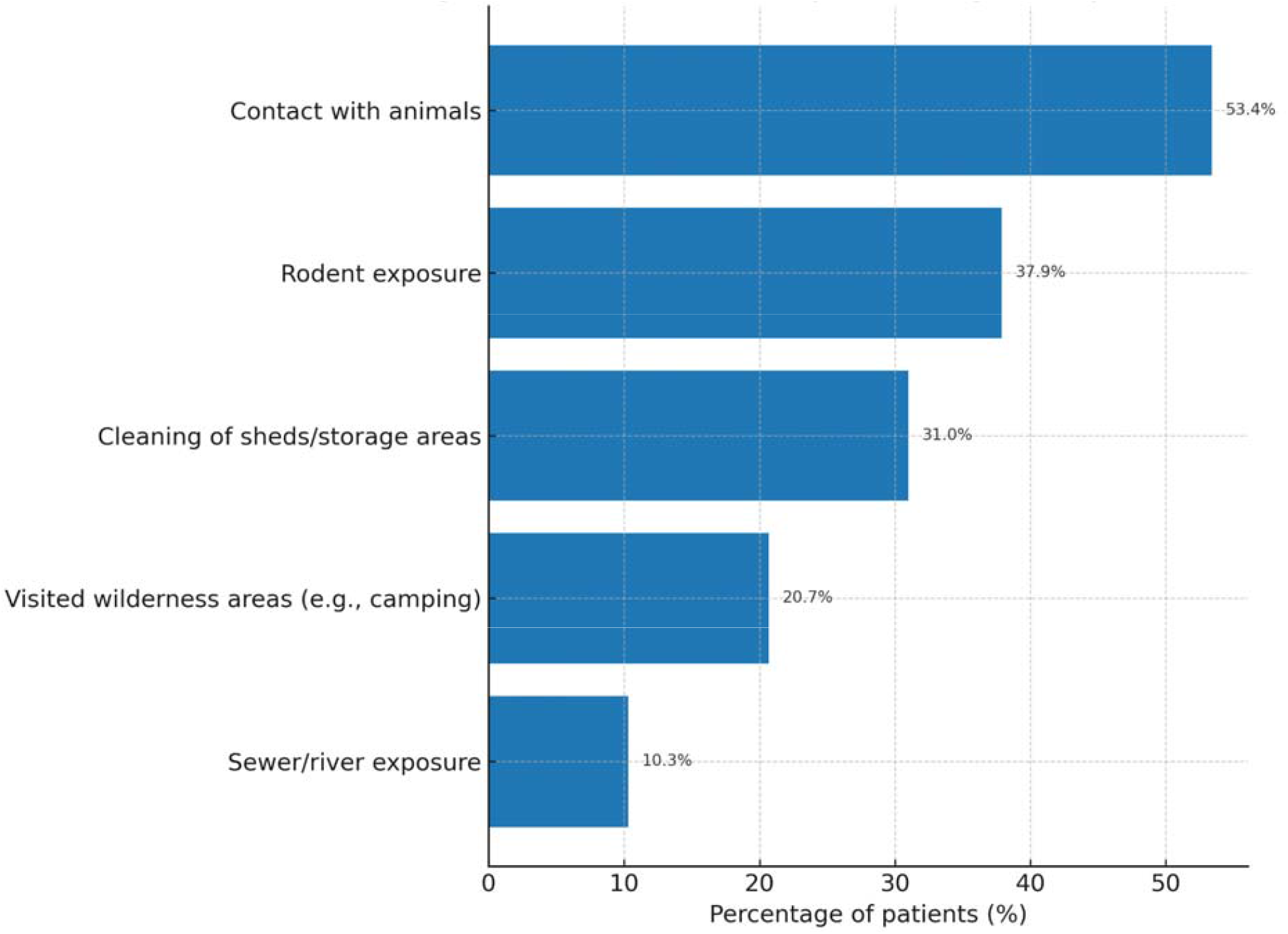
Environmental risk factors among Hantavirus cases, 2018-2021. Exposure within 30 days prior to symptom onset was recorded. Non-reported exposures were considered absent.

**Figure 4.**
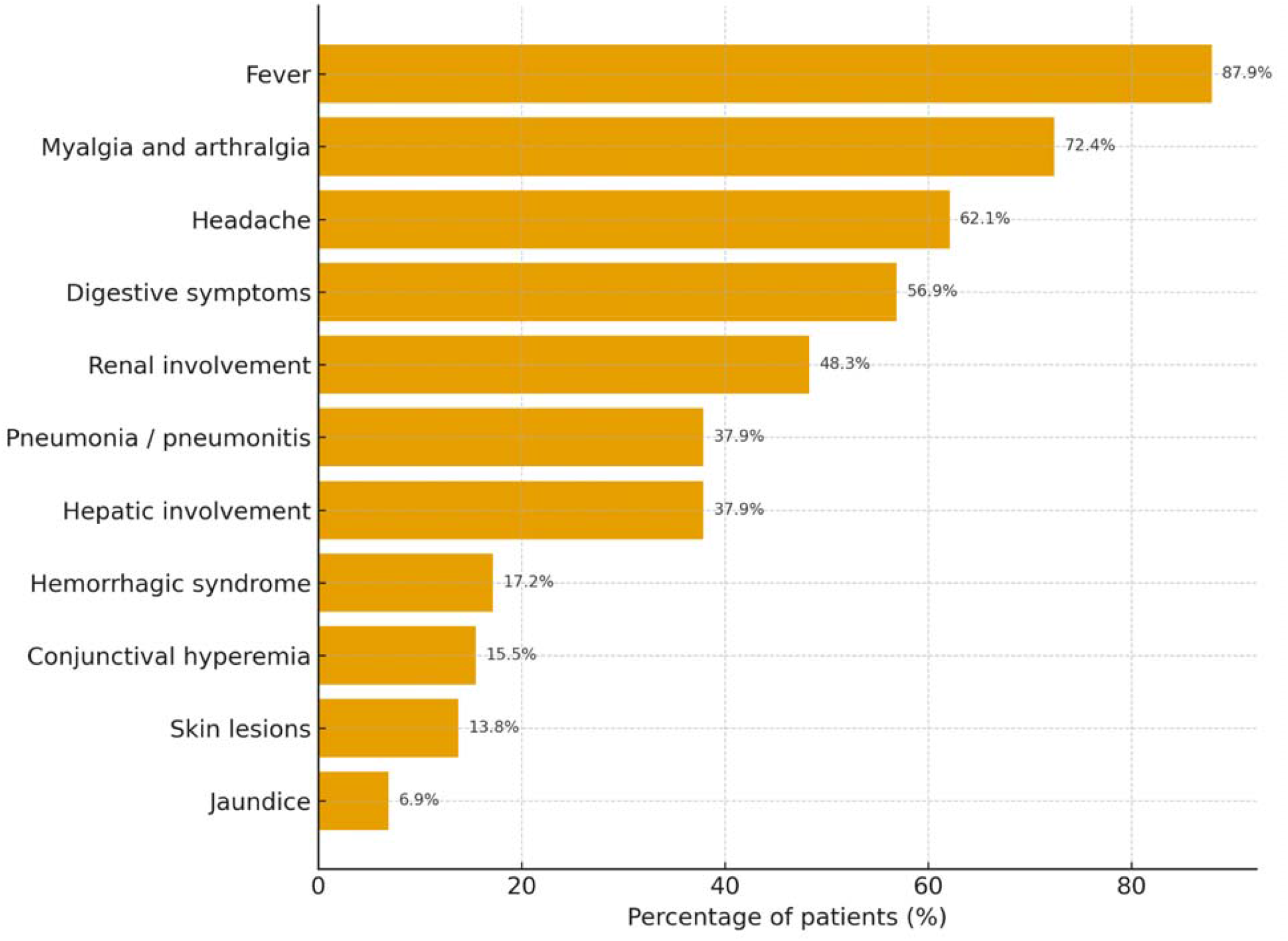
Distribution of clinical manifestations by frequency.

**Figure 5.**
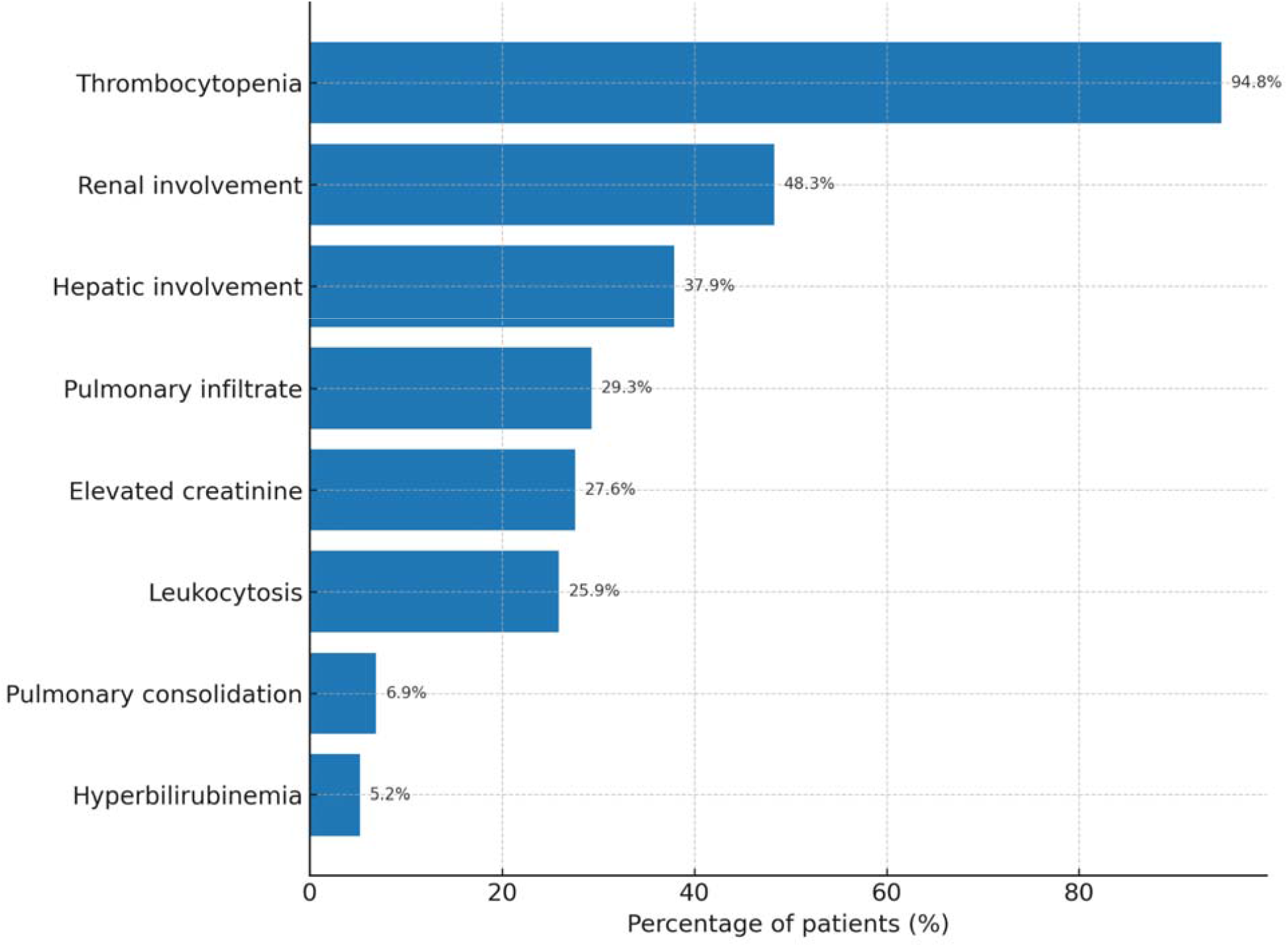
Frequency distribution of paraclinical results. Definitions: Thrombocytopenia = platelet count <150,000/mm^3^; Leukocytosis = white blood cell count >11,000/mm^3^; Elevated creatinine = >1.2 mg/dL; Hyperbilirubinemia = >1.2 mg/dL. Renal and hepatic involvement indicate clinical diagnosis documented in medical records. Missing laboratory values were considered within normal range.

### Associations with mortality

No significant association was found between mortality and biological sex. The highest proportion of deaths occurred in the 15-39 year age group (66.7%). Canelones had the highest case fatality rate (Figure 6). Fever, thrombocytopenia, and renal involvement were present in all fatal cases. Renal involvement was associated with a 3.4-fold increased odds of death (OR 4.4, 95% CI 1.04-19.01), and pneumonitis or pneumonia with a 4.5-fold increased odds (OR 4.5, 95% CI 1.12-17.93) (Table 2). No other clinical, demographic, or exposure variables were significantly associated with mortality. All but one fatal case had been hospitalized. (Table 2)

**Table 1.** Univariate analysis of factors associated with mortality in confirmed hantavirus cases.

| **Mortality-associated variable** | **OR** | **CI 95%** |
| --- | --- | --- |
| Sex (Male/Female) | 1.28 | 0.37 - 4.39 |
| Admission | 3.38 | 0.38 - 30.20 |
| Fever | 0.43 | 0.03 - 7.50 |
| Headache | 1.52 | 0.35 - 6.67 |
| Hemorrhage | 1.56 | 0.32 - 7.73 |
| Myalgia/Arthralgia | 1.71 | 0.32 - 9.30 |
| Renal failure | 4.44 | 1.04 - 19.02 |
| Pneumonia | 4.50 | 1.13 - 17.93 |
| Shed cleanup | 0.49 | 0.12 - 2.09 |
| Wilderness areas | 2.33 | 0.49 - 11.07 |
| Rodent exposure | 1.02 | 0.30 - 3.47 |
| Leukocytosis | 0.35 | 0.08 - 1.45 |
| Thrombocytopenia | 0.67 | 0.06 - 7.11 |
| Pulmonary consolidation | 2.60 | 0.28 - 23.81 |

CFR = (deaths/confirmed cases) × 100. Departments with 0 reported deaths were grouped as “Other (0% CFR)”. Values may be influenced by small sample sizes in some departments.

**Figure 6.**
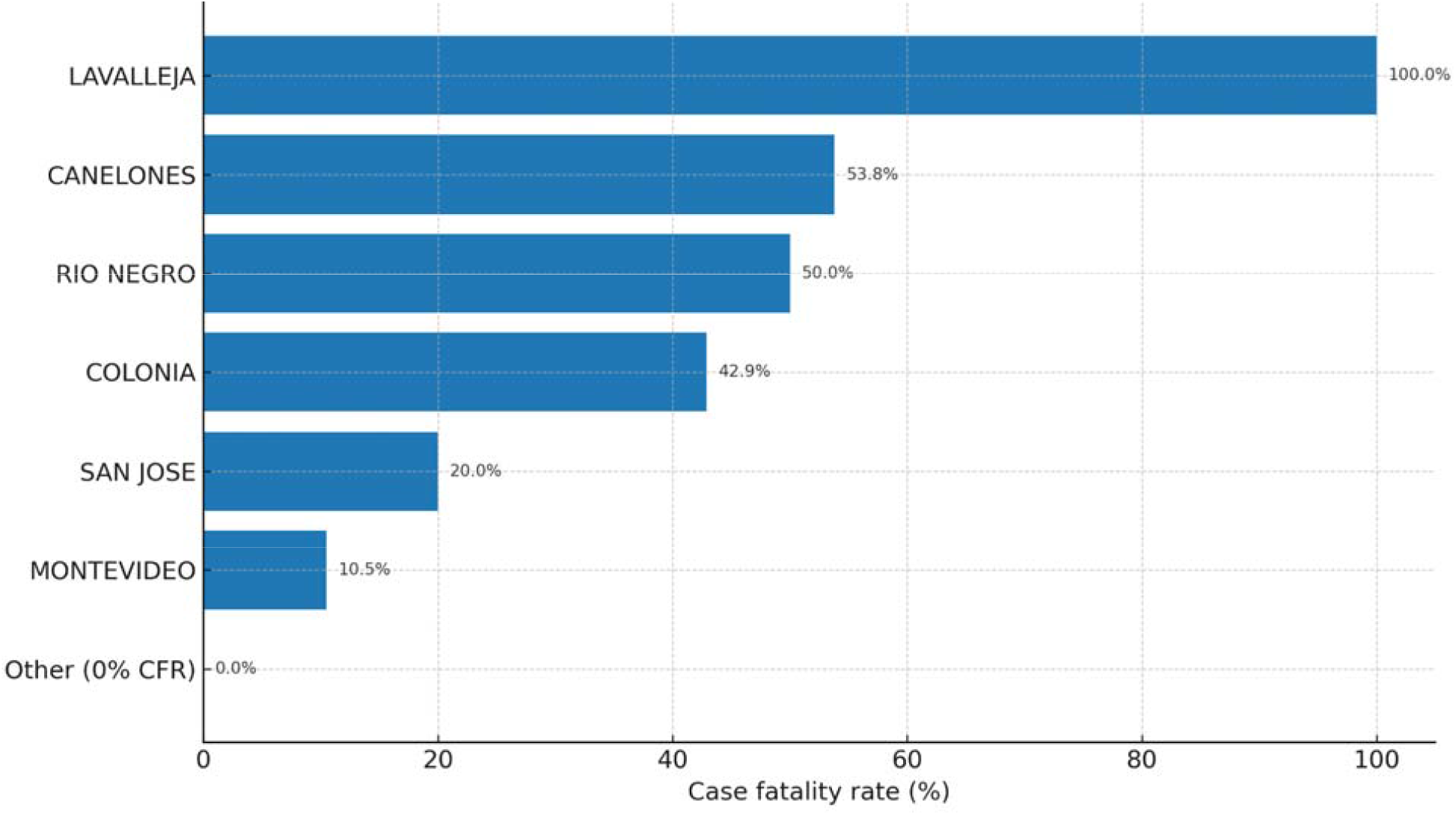
Case fatality rate (%) by department. CFR = (deaths/confirmed cases) × 100. Departments with 0 reported deaths were grouped as “Other (0% CFR)”. Values may be influenced by small sample sizes in some departments.

## Discussion

Hantavirus disease remains a significant public health concern in Uruguay, the wider region, and globally [4-8]. In our study, the decline in incidence observed during the SARS-CoV-2 pandemic years likely reflects multiple interacting factors, including reduced human mobility, widespread use of face masks, and a decrease in in-person medical consultations. These circumstances may have introduced a consultation bias, reducing case detection rather than true disease occurrence. Importantly, case fatality rates remained consistently high, underscoring the severity of clinically apparent infections.

Our analysis was limited to cases reported to the Ministry of Public Health (MPH), most of which were identified following in-person healthcare visits. This reliance on passive surveillance likely resulted in underreporting of mild or asymptomatic cases and an overrepresentation of severe presentations, potentially inflating case fatality estimates.

Consistent with previous reports, we found a predominance of cases in males, probably reflecting greater occupational exposure. The highest proportion of cases and deaths occurred in the 15–39-year age group, aligning with international evidence that hantavirus disease disproportionately affects young working populations [9].

Geographical patterns were also notable: most cases were concentrated south of the Río Negro, particularly in Montevideo and Canelones. This distribution mirrors findings from other endemic areas, where urban expansion increases human–rodent contact, and supports environmental hypotheses that conditions south of the Río Negro favor viral persistence. Broader environmental factors, including climate change and increased flooding, may further contribute to transmission dynamics [6].

Although delays in diagnosis could plausibly worsen outcomes, this could not be directly assessed in our dataset. Given that hantavirus disease often presents with nonspecific early symptoms and lacks a specific antiviral treatment, timely recognition and early initiation of supportive care remain critical to improving prognosis.

In light of the predominant inhalation route of infection, mandatory mask use during high-risk occupational activities should be considered as part of prevention strategies, reflecting one of the most important practical lessons learned during the COVID-19 pandemic.

Finally, despite the global relevance of hantavirus disease, its relatively low incidence has limited vaccine development interest. A vaccine (Hantavax®) is currently available in some Asian countries; however, it is not approved or available in Uruguay or in the region, and evidence regarding its efficacy remains limited [5,12]. Likewise, there is currently no specific antiviral therapy approved by the U.S. Food and Drug Administration (FDA); management remains supportive [10-12]. Nonetheless, recent advances in vaccine technology during the COVID-19 pandemic have demonstrated the potential for rapid innovation when resources and priorities align. A similar commitment could accelerate progress toward effective preventive measures against this neglected yet severe zoonosis, which disproportionately impacts the young working population.

## Conclusions

The incidence of hantavirus disease in Uruguay decreased between 2018 and 2021. When comparing the two years prior to the COVID-19 pandemic with the first two years of the pandemic, a marked decline in cases was observed; however, this was not accompanied by a significant reduction in the case fatality rate.

The vast majority of cases occurred south of the Río Negro, with Montevideo and Canelones being the most affected departments, likely related to increased urbanization, greater exposure to rodents, and environmental predisposition. The department with the highest case fatality rate was Canelones, followed by Colonia and Montevideo.

The most frequent symptoms among fatal cases were fever, myalgia and/or arthralgia, and headache, similar to other viral syndromes. The most common laboratory abnormalities were thrombocytopenia and renal failure. Renal and pulmonary involvement were the only variables independently associated with mortality.

These findings highlight the need for sustained epidemiological surveillance, early diagnosis, and targeted preventive measures, particularly in high-incidence urban areas south of the Río Negro in Uruguay.

## Data Availability

All data produced in the present study are available from the corresponding author upon reasonable request. Access to epidemiological surveillance data is subject to authorization by the Ministry of Public Health of Uruguay and applicable ethical and legal restrictions.

## Acknowledgments

We thank the Ministry of Public Health, Department of Epidemiological Surveillance; the Academic Medical Unit I, Hospital Maciel, Montevideo, Uruguay; and the Faculty of Medicine, Universidad de la República, for their collaboration in this research.

